# Antibody Response to COVID-19 vaccination in Patients Receiving Dialysis

**DOI:** 10.1101/2021.05.06.21256768

**Authors:** Shuchi Anand, Maria E. Montez-Rath, Jialin Han, Pablo Garcia, LinaCel Cadden, Patti Hunsader, Russell Kerschmann, Paul Beyer, Mary Dittrich, Geoffrey A Block, Scott D Boyd, Julie Parsonnet, Glenn M Chertow

## Abstract

**Background:** Patients receiving dialysis may mount impaired responses to COVID19 vaccination.

**Methods:** We report antibody response to vaccination from 1140 patients without, and 493 patients with pre-vaccination SARS-CoV-2 RBD antibody. We used commercially available assays (Siemens) to test remainder plasma monthly in association with vaccination date and type, and assess prevalence of absent total receptor binding antibody, and absent or attenuated (index value < 10) semiquantitative receptor binding domain IgG index values. We used Poisson regression to evaluate risk factors for absent or attenuated response to vaccination.

**Results:** Among patients who were seronegative versus seropositive before vaccination, 62% and 56% were ≥65 years old, 20% and 24% were Hispanic, and 22% and 23% were Black. Median IgG index values rose steadily over time, and were higher among the seropositive than in the seronegative patients after completing vaccination (150 [25^th^, 75^th^ percentile 23.2, 150.0] versus 41.6 [11.3, 150.0]). Among 610 patients who completed vaccination (assessed ≥14 days later, median 29 days later), the prevalence of absent total RBD response, and absent and attenuated semiquantitative IgG response was 4.4% (95% CI 3.1, 6.4%), 3.4% (2.4, 5.2%), and 14.3% (11.7, 17.3%) respectively. Risk factors for absent or attenuated response included longer vintage of end-stage kidney disease, and lower pre-vaccination serum albumin.

**Conclusions:** More than one in five patients receiving dialysis had evidence of an attenuated immune response to COVID19 vaccination.

**Significance statement:** Patients receiving dialysis face high likelihood of severe COVID19; at the same time, vaccination may be less efficacious, as prior data indicate impaired immune responses to influenza and Hepatitis B vaccination. We found that 22% of patients receiving dialysis had suboptimal responses to vaccination, irrespective of whether or not they had evidence of prior SARS-CoV-2 infection. Poorer health status and longer duration of end-stage kidney disease increased likelihood of suboptimal response. Ongoing vigilance for COVID19 in dialysis facilities and studies of modified vaccination dosing schedules will be critical to protecting patients receiving dialysis.

## Introduction

The COVID-19 pandemic has disproportionately affected patients on dialysis. Although SARS-CoV-2 seroprevalence among patients on dialysis is similar to the general population^1,2^, hospitalization rates have been 3-to-4 fold higher than other Medicare beneficiaries^3^. Nearly one-third of patients receiving dialysis died after hospitalization with COVID-19 in New York^4^, Spain^5^ and Italy^6^, a 20-30 fold higher mortality rate compared with the general population. More recent data continue to show mortality rates in this population in excess of 15%^7^.

As of March 2021, dialysis centers have received direct allotments of COVID-19 vaccines. A majority (80%) of patients receiving hemodialysis expressed willingness to get vaccinated^8^. Whether vaccination will result in efficacy similar to that seen in the general population, however, is yet unknown. In influenza vaccine studies—one focused on H1N1 strain alone^9^ and one^10^ on trivalent vaccine including H1N1 strain—57% and 46% of patients receiving dialysis respectively mounted immunologically significant titers to H1N1 at 4 weeks, compared with 90% or more of healthy volunteers. Data from hepatitis B^11^ and influenza vaccination^9,10,12^ provides incontrovertible evidence of blunted and foreshortened response to immunization in patients receiving dialysis^11,13-16^, raising the worrisome possibility that the SARS-CoV-2 vaccine may also have lower efficacy among patients on dialysis.

In order to assess response to SARS-CoV-2 vaccination among patients on dialysis, we have been following 4,346 patients on dialysis without prior SARS-CoV-2 antibody since January 2021. Since August 2020, we have followed 6551 patients among whom a subset seroconverted prior to vaccination (i.e., developed evidence of natural infection). In this study, we report early data on receptor binding domain (RBD) seroconversion and semi-quantitative IgG values post vaccination in 1140 patients without, and 493 patients with, pre-vaccination SARS-CoV-2 RBD antibody. Among the subset who completed two doses of vaccination, we assess rates of and risk factors for absent or attenuated response. We hypothesized that patients of poorer health status—i.e., older patients, patients of longer end-stage kidney disease (ESKD) vintage, and those with lower serum albumin—would be more likely to exhibit an absent or attenuated response.

## Methods

Our study was conducted in partnership with US Renal Care, the third largest dialysis network in the U.S., and Ascend Clinical Laboratory, which processes monthly laboratories of patients receiving dialysis at US Renal Care and other facilities. Ascend Clinical tested remainder plasma of patients for SARS-CoV-2 antibody, and anonymized all patient demographic, comorbidity, and laboratory data prior to transfer to Stanford University. The Institutional Review Board at Stanford University reviewed and approved the study.

Sample size: In the first two weeks of January 2021—prior to widespread vaccine roll out—we tested SARS-CoV-2 antibody status of 21,570 patients on hemodialysis. From among the 17,390 seronegative patients in January 2021, we used systematic sampling with fraction intervals stratified by age to randomly select 4,346 persons to follow with monthly SARS-CoV-2 serology assays, in association with type and date of vaccination(s) (**Figure 1**). Since our primary goal was to describe immune responses to vaccine, and since we hypothesized that older patients would be less likely to mount an immune response than younger patients, we estimated the sample size using age-stratified response rates to hepatitis B vaccination (see **Supplemental Appendix 1** for details).

**Figure 1.**
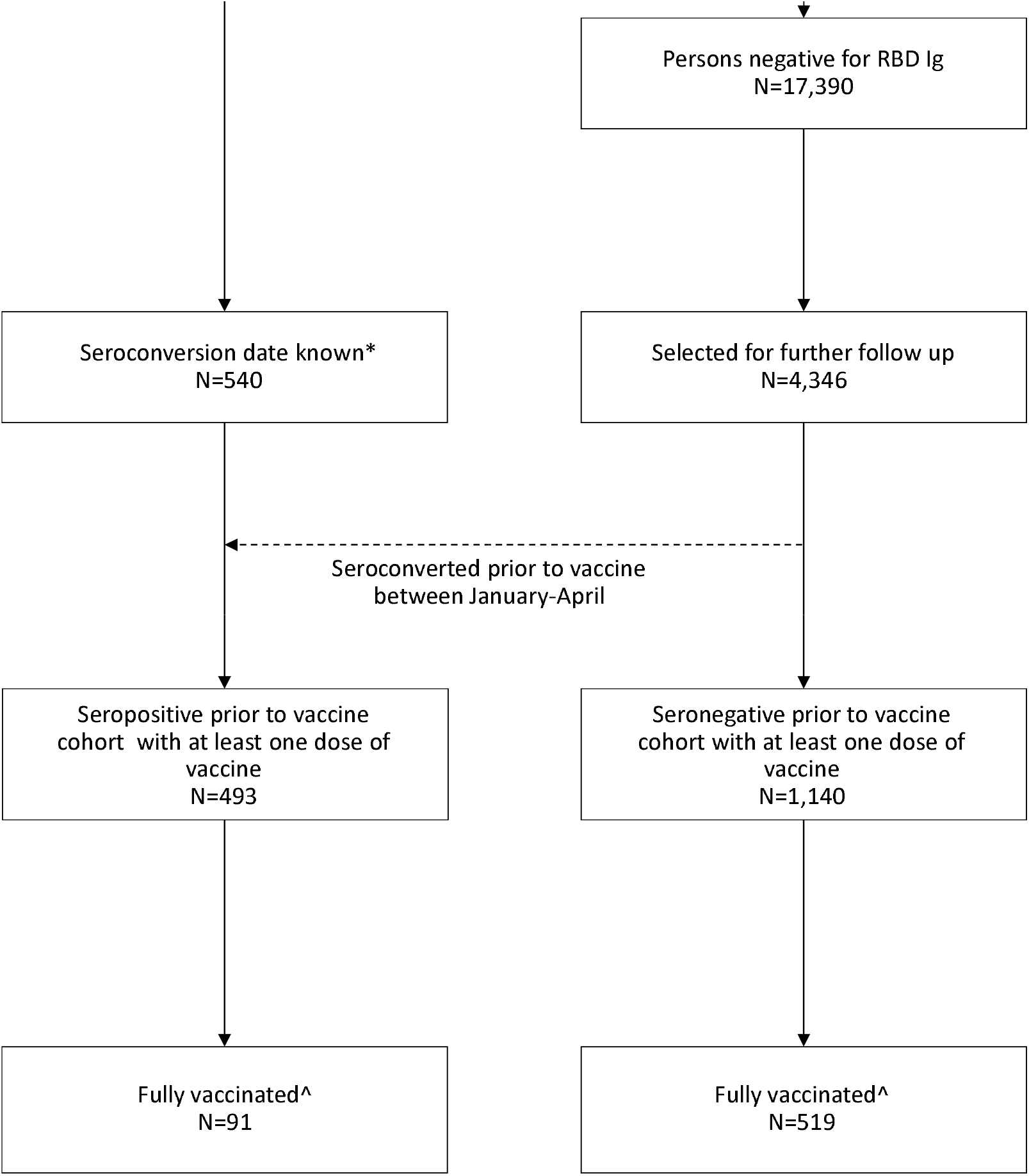
Flow chart of study cohorts

All 540 patients seropositive as of January 2021, and any additional patients who seroconverted prior to vaccination (**Figure 1**) comprised the “seropositive prior to vaccination cohort”. We can approximate the dates of seroconversion in all participants since we have followed them monthly since July of 2020. The participants without SARS-CoV-2 antibodies prior to vaccination comprise the “seronegative prior to vaccination” cohort.

### Assay Characteristics

We tested remainder samples using the Siemens’ total RBD Ig assay, which measures IgG and IgM antibodies, in January 2021 and monthly thereafter in the seronegative prior to vaccination cohort. This assay is reported by the manufacture to have 100% sensitivity and 99.8% specificity for tests performed ≥14 days after a positive reverse transcriptase polymerase chain reaction test^17^; it has been validated independently with similar performance characteristics^18,19^. Subsequent to a positive total RBD Ig result in the seronegative and among all patients in the seropositive prior to vaccination cohorts, we tested samples only using a semiquantitative Siemens RBD IgG assay monthly. The Siemens RBD IgG assay is semiquantitative two-step sandwich indirect chemiluminescent assay with a manufacturer reported 95.6% (95% CI: 92.2-97.8%) sensitivity and 99.9% (95% CI 99.6-99.9%) specificity for tests performed ≥21 days post positive reverse transcriptase polymerase chain reaction test. An index value ≥1.0 is considered reactive and an index value of 150 is the upper limit of quantification.

### Evaluation of Antibody Responses

We evaluated response over three time periods: <14 days post first dose of vaccine (early post-vaccine), between 14 days post first dose and 14 days post second dose (partially vaccinated, applicable only to the mRNA platform vaccines), and > 14 days post the Johnson and Johnson vaccine or second dose of the mRNA platform vaccines or (fully vaccinated). If multiple samples were available within each period, we selected the IgG result earliest in that period. Among fully vaccinated patients, for example, this represents the IgG result closest to 14 days post vaccination. We classified responses as absent total RBD Ig antibody, absent semiquantitative IgG antibody (index value <1), attenuated antibody (semiquantitative IgG index value <10) or medium to high antibody (≥10)^20^. We chose 10 as a cut-point based on data showing that index values ≥10 corresponded with pseudovirus neutralization titers^21^ > 1:500 in a study of 26 patients tested by Siemens. An additional study (n=74) evaluating correlation with plaque reduction neutralization test reported index values ≥10 had a positive predictive value of 100% for plaque reduction neutralization test_50_ >1:80 ^22,23^. Finally, in an external study of patients with inflammatory bowel disease on biologic therapy post SARS-CoV-2 vaccination, 22 of 26 patients with inflammatory bowel disease and all 14 healthcare workers who had completed vaccination exhibited semiquantitive titers with index values > 10^24^.

### Correlates

We extracted electronic health record data on age, sex, self-reported race and ethnicity, years with end-stage kidney disease, diabetes status, and nursing home status as available. We also extracted monthly laboratory results for serum albumin—a valid surrogate of health status^25-28^. We used the laboratory value closest to the date prior to vaccination.

### Statistical analysis

We present demographic data and laboratory values using proportions, mean ± standard deviation (SD) or median, 25^th^-75^th^ percentile, as applicable. We present the range of semiquantitative IgG titers by vaccine period in the seronegative and seropositive prior to vaccination cohorts. Among patients in the ‘fully vaccinated’ window, we present prevalence and 95% confidence intervals, overall and by age group, of absent or attenuated antibody response in the overall, and seronegative and seropositive prior to vaccination cohorts. Finally, we present these parameters by vaccine type. In a sensitivity analysis, we assessed prevalence of absent or attenuated antibody response after at least 28 days post completion of vaccination. Among participants who completed vaccination, we used a Poisson model with robust standard error to assess risk factors for absent or attenuated antibody response. Data missingness was low (5%), and exclusively due to missing self-reported race/ethnicity. We therefore present results of a complete case analysis inclusive of both cohorts, in which we *a priori* selected the following correlates to test: age, sex, race/ethnicity, diabetes status, vintage of ESKD, and serum albumin. In a second model, we tested whether age or serum albumin modified the association between seropositive status prior to vaccination and the post-vaccination antibody response.

We considered statistical significance at α<0.05. We conducted all statistical analyses with SAS (Cary, North Carolina) or Stata/MP 16.1 (College Station, TX 2019).

## Results

As of April 2021, 1140 patients on dialysis without prior SARS-CoV-2 antibodies and 493 patients with extant antibodies received a dose of vaccine (**Figure 1**). **Table 1** shows demographic and clinical characteristics of the seronegative and seropositive before vaccination cohorts.

**Table 1.**
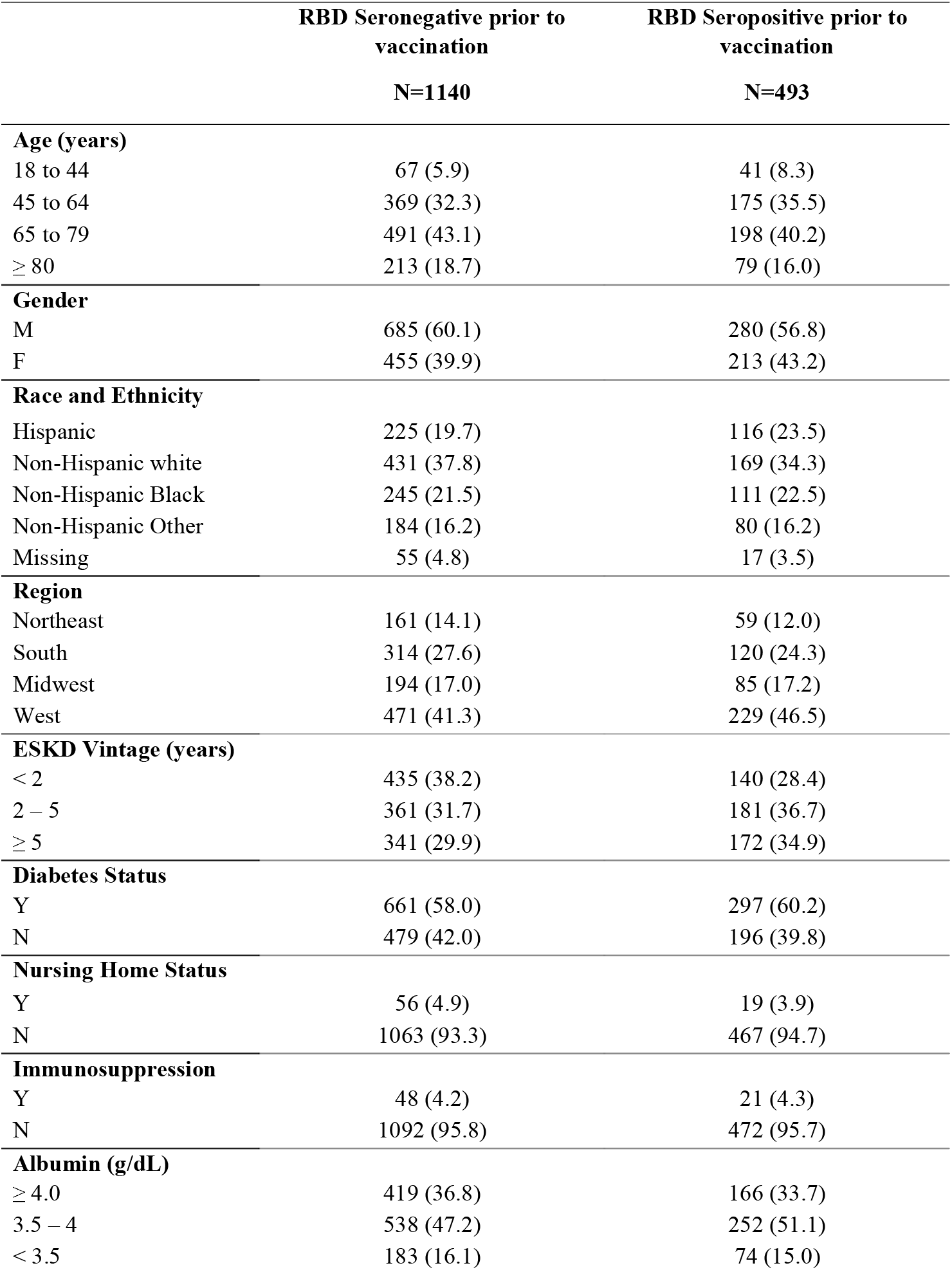
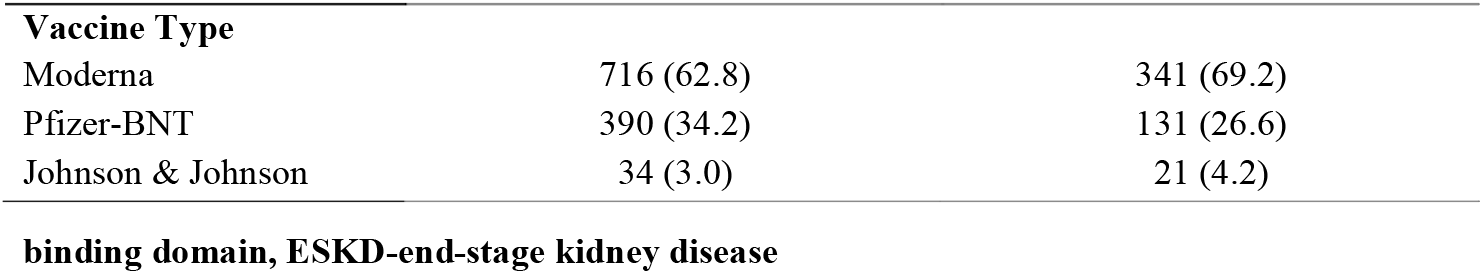
Participant characteristics according to SARS-CoV-2 spike protein receptor binding domain antibody status prior to vaccination

In the seronegative prior to vaccination cohort, 9%, 35%, and 77% of patients had RBD IgG values ≥10 in the early post-vaccine, partially vaccinated, and fully vaccinated groups, respectively. In the seropositive cohort, the proportions were higher in the earlier periods (38%, 55%, and 81%, respectively). Median IgG levels were lower in the seronegative compared with the seropositive cohorts, starting at 14 days after the first dose (**Figure 2**).

**Figure 2.**
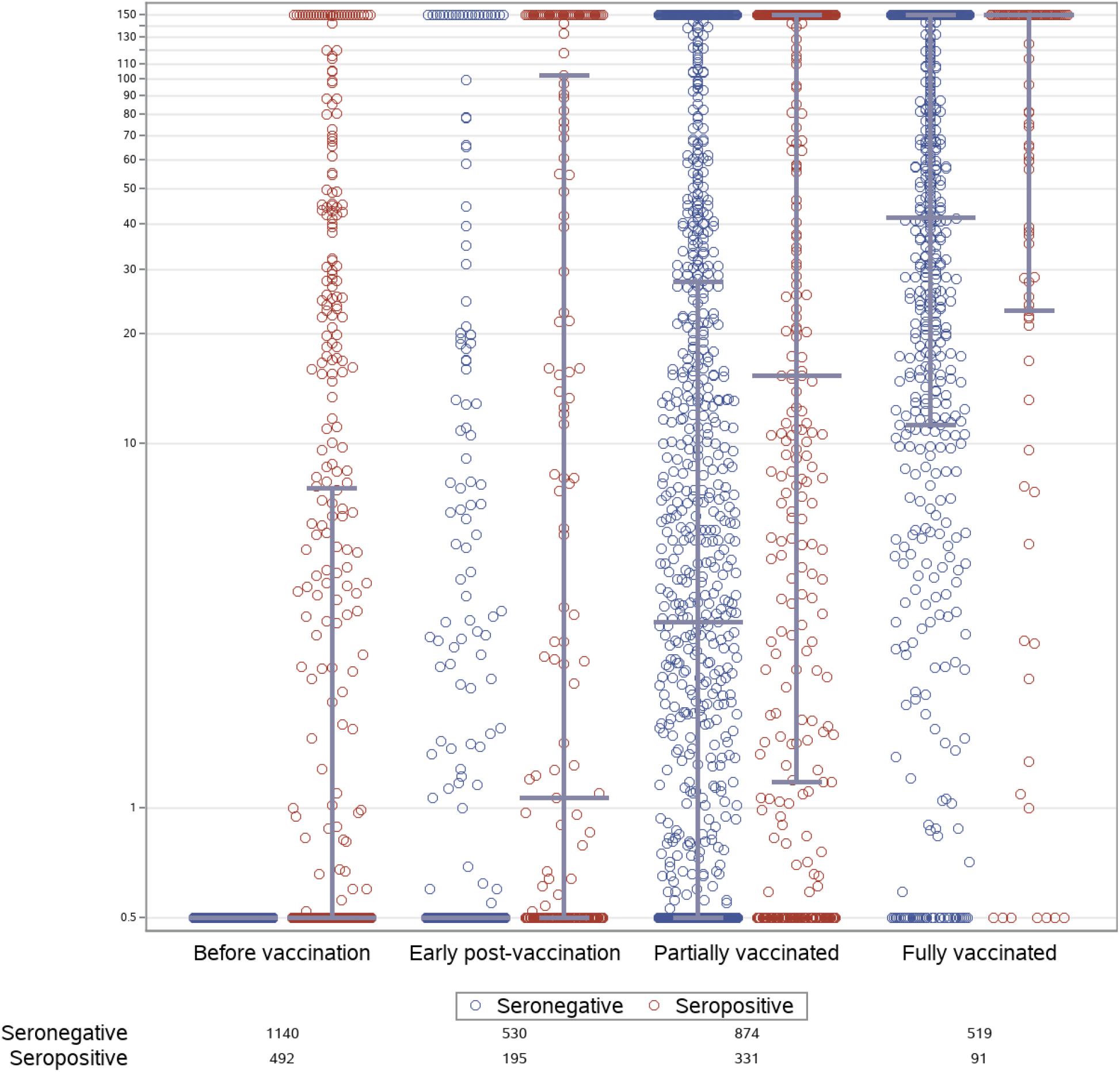
IgG responses following COVID19 vaccination among patients receiving dialysis Individual semiquantitative RBD IgG index values are graphed by time period since vaccination; the lines represent median with interquartile range. We defined early post vaccination as within 14 days of first dose, partially vaccinated as between 14 days after first dose to 14 days post second dose, and fully vaccinated as more than 14 days post second dose for Moderna and Pfizer. For Johnson and Johnson, we defined fully vaccinated as more than 14 days post first dose. In the seropositive prior to vaccination cohort, IgG index values closest to the dates before vaccination are graphed in the ‘before vaccination’ period; the seronegative cohort prior to vaccination was tested only with the total RBD Ig and all had negative results in the ‘before vaccination’ period. Although 1140 and 493 patients were included in the two cohorts (since they received at least one vaccine dose at the time of the study), all patients do not have values available for each time period, since these depend on the timing of the vaccination as well as on the timing of the routine monthly blood draw. For both cohorts, median IgG levels rose steadily after vaccination. Among the seronegative prior to vaccine cohort, median IgG index values post vaccination were 0.5 (25^th^, 75^th^ percentile 0.5, 0.5), 3.2 (0.5, 27.8), and 41.6 (11.3, 150.0) in the early, partially and fully vaccinated periods respectively. Among the seropositive prior to vaccine cohorts, median IgG index values post vaccination were 0.5 (0.5, 7.6), 1.1 (0.5, 102.3), 15.3 (1.2, 150), and 150 (23.2, 150) in the prior to, early, partially and fully vaccinated periods respectively.

We assessed vaccine response among fully vaccinated patients a median of 29 days (25^th^, 75^th^ percentile 22, 39) days post vaccine completion. Of the 610 fully-vaccinated patients, 27 (estimated prevalence 4.4% [95% CI 3. 1, 6.4%]), 21 (3.4% [2.4, 5.2%]), and 87 (14.3% [11.7, 17.3%]) had absent total RBD, absent semiquantitive IgG, and attenuated IgG response, respectively. The prevalence of absent or attenuated response was similar in seronegative and seropositive cohorts (**Table 2**). In both cohorts, there was a higher prevalence of an attenuated response in older age groups, but the trend was not linear. In a sensitivity analysis assessing responses at least 28 or more days post vaccine completion, there was no difference in the prevalence of absent-to-attenuated responses in seronegative and seropositive cohorts, overall or by age categories (**Supplemental Table 2**).

**Table 2.** Prevalence of absent or attenuated response among fully vaccinated individuals overall and by age group, at least 14 days after completion of vaccine*

|  | Seronegative prior to vaccination cohort |  |  | Seropositive prior to vaccination |  |
| --- | --- | --- | --- | --- | --- |
|  | N=519 |  |  | N=91 |  |
|  | No seroconversion on total RBD Ig | No detectable response on RBD IgG | Attenuated IgG | No detectable response on RBD IgG | Attenuated IgG |
| Age (years) |  |  |  |  |  |
| 18 to 44 | 9.1% (2.8, 30) | 0% (0, 0) | 4.5% (0.6, 26.2) | 0% (0, 0) | 0% (0, 0) |
| 45 to 64 | 6.8% (3.5, 13.1) | 0.9% (0.1, 5.8) | 9.4% (5.3, 16.2) | 5.6% (0.8, 31.2) | 5.6% (0.8, 31.2) |
| 65 to 79 | 5.5% (3.2, 9.3) | 3.8% (2.0, 7.2) | 16.5% (12.3, 21.8) | 8.3% (3.1, 20.4) | 12.5% (5.7, 25.4) |
| ≥ 80 | 2.8% (1.0, 7.2) | 2.8% (1.0, 7.2) | 18.1% (12.6, 25.2) | 8.7% (2.1, 29.3) | 13.0% (4.2, 33.9) |
| **Overall** | **5.2% (3.6, 7.5)** | **2.7% (1.6, 4.5)** | **14.8% (12.0, 18.2)** | **7.7% (3.7, 15.4)** | **11.9% (6.0, 19.2)** |

\*Data are percentage (95% CI) obtained at least 14 days after two doses of either Moderna or Pfizer-BNT vaccines and 14 days after a single dose of Johnson & Johnson vaccine. Median duration since completion of vaccination was 29 days [25<sup>th</sup>, 75<sup>th</sup> percentile: 22, 39 days]. We performed total RBD Ig among all patients in the seronegative prior to vaccination cohort; once a patient seroconverted, we performed the semiquantitative RBD IgG monthly. We performed semiquantitative RBG IgG only among patients known to have a positive total RBD Ig prior to vaccination (seropositive prior to vaccination cohort).

Non-white race and Hispanic ethnicity were associated with a lower risk of absent or attenuated response; longer ESKD vintage and lower serum albumin were associated with a higher risk (**Table 3**). Age did not modify the association between pre-vaccination antibody status and post-vaccination response (p-value for interaction with age 0.7704). There was a suggestion that serum albumin might modify the association between pre-vaccination antibody status and post-vaccination response (p-value for interaction 0.0546). Fourteen of 48 fully-vaccinated patients living in nursing homes (29%) and 13 of 27 (48%) on immunosuppressive medications had absent or attenuated response.

**Table 3.** Risk factors for absent or attentuated response to SARS-CoV-2 vaccination in fully vaccinated patients receiving dialysis

|  | Risk Ratio<sup>^</sup> |
| --- | --- |
| **Age (years)** |  |
| <65 | Ref |
| ≥65 | 1.22 (0.81-1.85) |
| **Race and ethnicity** |  |
| Non-Hispanic white | Ref |
| Hispanic | 0.37 (0.21-0.65) |
| Non-Hispanic Black | 0.88 (0.60-1.31) |
| Non-Hispanic Other | 0.61 (0.38-0.98) |
| **ESKD Vintage (years)** |  |
| < 2 | Ref |
| 2 to <5 | 1.39 (0.96-1.99) |
| ≥ 5 | 1.62 (1.11-2.36) |
| **Diabetes** |  |
| No | Ref |
| Yes | 0.90 (0.66-1.23) |
| **Serum albumin (0.5 g/dL) change** | 0.77 (0.63-0.93) |

<sup>^</sup>Includes seronegative and seropositive cohorts; adjusted for all presented correlates

In evaluating semiquantitative titers by vaccine type, median titers were modestly lower for Pfizer than for the Moderna vaccine (**Supplemental Figure 1a & b**), and correspondingly, there was a modestly higher prevalence of absent or attenuated response in the subgroup receiving Pfizer compared with Moderna (**Table 4**). Data on the disabled adenovirus Johnson and Johnson vaccination were sparse, but suggested higher prevalence of absent response among patients evaluated at 14 days post vaccination (**Table 4**).

**Table 4.** Prevalence of absent or attenuated response among fully vaccinated individuals by vaccine type, at least 14 days after completion of vaccine*

| Fully vaccinated |  |  |  |
| --- | --- | --- | --- |
| N=610 |  |  |  |
|  |  | No detectable<br>response on total RBD<br>or RBD IgG | Attenuated IgG |
| Moderna | 353 | 2.8% (1.5, 5.2) | 9.1% (6.5, 12.5) |
| Pfizer-BNT | 239 | 9.6% (6.5, 14.1) | 22.6% (17.7, 28.3) |
| Johnson & Johnson | 18 | 83.3% (59.1, 94.5) | 5.6% (0.8, 30.7) |

\*Data are percentage (95% CI) obtained at least 14 days after two full doses of either Moderna or Pfizer-BNT vaccines and 14 days after a single dose of Johnson & Johnson vaccine. We performed total RBD Ig among all patients in the seronegative prior to vaccination cohort; once a patient seroconverted, we performed the semiquantitative RBD IgG monthly. We performed semiquantitative RBG IgG only among patients known to have a positive total RBD Ig prior to vaccination (seropositive prior to vaccination cohort).

## Discussion

In our study of the early response to SARS-CoV-2 vaccination among patients on dialysis, we find that 8% have absent and 14% have attenuated RBD IgG response when evaluated at least 14 days after completing vaccination. Our data are in line with some recently published reports^29,30^, and portend the critical need for studies evaluating real-world efficacy of vaccination in this and other vulnerable populations with chronic diseases, and for trials evaluating modified schedules of vaccination. We also found that, although median IgG titers are higher among patients with evidence of prior SARS-CoV-2 infection compared with those without, rates of absent- or attenuated response to vaccination were similar between the two groups. There were differences in responses by vaccine type that require further study, since our cohort is small and patient characteristics may have influenced type of vaccine delivered. Longer ESKD vintage and lower serum albumin concentrations were associated with an attenuated response to SARS-CoV-2 vaccination.

Several small studies have recently supplied data on antibody response to vaccination in patients with ESKD^29-31^ and other chronic diseases. In a study of 56 patients on dialysis and 95 healthcare workers from Israel also measuring semi-quantitative IgG responses a median of 30 days post second vaccination, the authors found a lower prevalence of non-response (4%) and noted lower median semi-quantitive titers in the patients receiving hemodialysis (2900 AU/ml) compared with healthcare workers (7400 AU/mL)^29^. Canaday *et al*. report 50% lower anti-spike and anti-RBD titers and 4-fold lower neutralizing antibody titers among nursing home residents compared with health care workers^32^. Concordantly, our study indicates a worrisome prevalence of attenuated response among patients on dialysis post vaccination, especially in the context of data from health care workers showing that 100% had evidence of RBD seroconversion and of IgG index values >10 post vaccination^24^, and our data indicating that a majority of patients surviving SARS-CoV-2 infection had IgG index values > 10^20^.

Some countries have prioritized rapid dissemination of the first dose of the vaccine to as many people as possible over ensuring a second dose is delivered per schedule to a smaller population^33,34^. Our data indicating that more than half of vaccinated patients on hemodialysis remain at RBD IgG index values <10 more than 14 days after their first dose of the mRNA vaccines strengthen the rationale for providing a second dose to this population as per clinical trial informed schedules. Our findings also suggest that a third or ‘booster’ dose may be necessary to optimally protect patients on dialysis. Patients with prior evidence of SARS-CoV-2 infection reached higher RBD IgG, more comparable to those reported in the general population^24^. Thus, it is conceivable that modified (3-dose) vaccination schedule would yield better efficacy as measured by antibody titers. A certain fraction may nonetheless remain vulnerable to infection or reinfection despite modified vaccination schedules, as suggested by the ∼20% prevalence of absent or attenuated response among fully vaccinated patients with prior evidence of infection in our study. Even as community-wide mitigation strategies are relaxed in response to widespread vaccination, ongoing surveillance and testing in dialysis facilities may be required.

Our data signal a lower likelihood of a measurable antibody response to vaccination in the small subgroup of patients receiving the disabled adenovirus vaccine. However, these results require cautious interpretation, since the number of patients evaluated was small (n=55), and since the allocation of vaccines was not randomized. The modest differences in IgG index values and likelihood of a attenuated antibody response between the mRNA platform vaccines have been confirmed in other populations^35^. Further studies are required to determine whether these differences translate to differences in the likelihood of serious COVID19 illness in populations with higher levels of comorbidity, frailty or impaired immunity than those enrolled in clinical trials assessing vaccine safety and efficacy.

Not surprisingly, we found that patients with lower serum albumin were more likely to have absent or attenuated responses^36^. Age was not associated with the likelihood of absent or attenuated response in our study, although the modest sample size and selection effects does not rule out an association between older age and an attenuated immune response to vaccination. Longer dialysis vintage was associated with a higher risk for absent or attenuated response, suggesting that the cumulative effects of ESKD (e.g., reflected by chronic inflammation, malnutrition, sarcopenia, and/or frailty) may diminish adaptive immune responses, as has been suggested by Kato *et al*.^37^ and others^38,39^. Black, Hispanic and other racial minority patients experienced substantially lower risks for absent to attenuated response, consistent with data on improved survival on dialysis in these subgroups^40^. Higher efficacy as assessed by antibody response may provide an additional data point to assist nephrologists in facilitating vaccine uptake.

Strengths of our study include use of a sensitive and specific commercially available assay which can be used on a large-scale to evaluate response to vaccination, the inclusion of a cohort with evidence of prior SARS-CoV-2 infection, and detailed data on patient health status. We measured immunologic response to RBD, one of the major criteria by which phase I data on the mRNA platform vaccines were evaluated.

Limitations of the study include the modest sample size. Our assessment was performed during the early phase of vaccine roll out, a time period during which elderly or persons with comorbidities were prioritized. Estimates for vaccine response may improve over time as a broader patient population receives vaccination, although 40% of our cohort was < 65 years of age indicating reasonable representativeness by age of patients receiving dialysis. Antibody titers are only one way to assess immunologic response to vaccination. We do not yet know whether the strength or duration of a measurable antibody response correlates with protection from infection.

In summary, in a well-characterized cohort of patients receiving dialysis with and without prior evidence of infection with SARS-CoV-2, more than one in five demonstrated an attenuated immune response after vaccination with one of three vaccines granted emergency use authorization by FDA. These findings may inform subsequent vaccination strategies in the ESKD population and in other persons with chronic illness.

## Data Availability

Requests for data will be reviewed by the authors, and data made available on a case-by-case basis.

