## Supplemental Tables1&2, Figure1 for "Antibody Response to COVID-19 vaccination in Patients Receiving Dialysis"

**Table of Contents**

**Appendix:** Sampling Strategy	p1

**Supplemental Table 1.** Population and subpopulation sizes by age and number of patients required to obtain a prevalence estimate with the specified absolute precision assuming and the specified proportion of non-response to the vaccine.	p2

**Supplemental Table 2.** Prevalence of absent or attenuated response among fully vaccinated individuals overall and by age group, at least 28 days after completion of vaccine	p3

**Supplemental Figure 1a&b:** Semiquantitative IgG values in patients receiving Moderna (a) or Pfizer (b)

p4

**Appendix:** Sampling Strategy

We selected 4348 patients on follow from the 17,390 patients on dialysis without prior evidence of SARS-CoV-2 infection (as of January 2021) in US Renal Care network. To estimate sample size, we used previously published data on hepatitis B vaccination non-response, as available by age strata from Bruguera et al., who evaluated immune response in 270 patients. In this study, the rate of non-response among persons age 20-40, 40-60, and > 60 years was 7%, 13%, and 35% respectively. Correspondingly, our estimates of non-response among persons age 18-44, 45-64, ≥65 years was 5%, 15%, and 30% respectively. Estimating these proportions of non-response with an absolute precision of 2%, and oversampling by 15%, resulted in a sample size estimate of 4222 (Supplemental Table 1).

**Supplemental Table 1**. Population and subpopulation sizes by age and number of patients required to obtain a prevalence estimate with the specified absolute precision assuming and the specified proportion of non-response to the vaccine.

| **Age group** | **Proportion of non-response to vaccine** | **Absolute precision** | **USRDS Population count** | **US Renal Care Population size** | **Sample size required** | **Over sample (15%)** |
| --- | --- | --- | --- | --- | --- | --- |
| 18 to 44 | 5% | 2% | 60,540 | 2,871 | 453 | 521 |
| 45 to 64 | 15% | 2% | 207,022 | 10,605 | 1,218 | 1,401 |
| ≥ 65 | 30% | 2% | 231,588 | 12,777 | 2,000 | 2,300 |
| Total |  |  | 499,150 | 26,253 | 3,671 | 4,222 |

We used systematic sampling with fractional intervals. In systematic sampling the patients are selected from the list using a fixed selection interval, calculated by dividing the total number of patients in the list by the desired number (i.e., 17390/4222 = 4.1). We thus randomly selected one number between 1 and 4 and then selected every 4th patient in the sampling frame sorted by zip code, age, sex and race. This resulted in a sample size of 4348 patients on dialysis; however 2 of sampled patients seroconverted prior to vaccination, thus we followed 4346 patients from January 2021.

|  | Seronegative prior to vaccination cohort  N=355 | | |  | Seropositive prior to vaccination  N=48 | |
| --- | --- | --- | --- | --- | --- | --- |
|  | No seroconversion on total RBD Ig | No detectable response on RBD IgG | Attenuated IgG |  | No detectable response on RBD IgG | Attenuated IgG |
| Age (years) |  |  |  |  |  |  |
| 18 to 44 | 7.1% (1, 37.2) | 0% (0, 0) | 0% (0, 0) |  | 0% (0, 0) | 0% (0, 0) |
| 45 to 64 | 1.4% (0.2, 9) | 0% (0, 0) | 14.9% (8.4, 24.9) |  | 0% (0, 0) | 10% (1.3, 48.1) |
| 65 to 79 | 3.8% (1.7, 8.2) | 3.1% (1.3, 7.4) | 24.5% (18.4, 31.8) |  | 13% (4.1, 34.3) | 17.4% (6.5, 38.9) |
| ≥ 80 | 1.9% (0.5, 7.1) | 4.6% (1.9, 10.7) | 25.0% (17.7, 34.0) |  | 0% (0, 0) | 14.3% (3.5, 43.7) |
| **Overall** | **2.5% (1.3, 4.6)** | **3.2% (1.9, 5.5)** | **20.8% (17.1, 25.1)** |  | **6.3% (2.0, 18.1)** | **14.6% (7.0, 28.0)** |

**Supplemental Table 2.** Prevalence of absent or attenuated response among fully vaccinated individuals overall and by age group, at least 28 days after completion of vaccine*

*Data are percentage (95% CI) obtained at least 28 days after two full doses of either Moderna or Pfizer-BNT vaccines and 28 days after a single dose of Johnson & Johnson vaccine. We performed total RBD Ig among all patients in the seronegative prior to vaccination cohort; once a patient seroconverted, we performed the semiquantitative RBD IgG monthly. We performed semiquantitative RBG IgG only among patients known to have a positive total RBD Ig prior to vaccination (seropositive prior to vaccination cohort). Among both cohorts the prevalence of no seroconversion on total RBD Ig, no detectable response on RBD IgG and attenuated IgG was 2.8% (1.5, 5.2), 2.8% (1.5, 5.2) and 72.7 (67.8, 77.1) respectively.

**Supplemental Figure 1a&b:** Semiquantitative IgG values in patients receiving Moderna (a) or Pfizer (b)

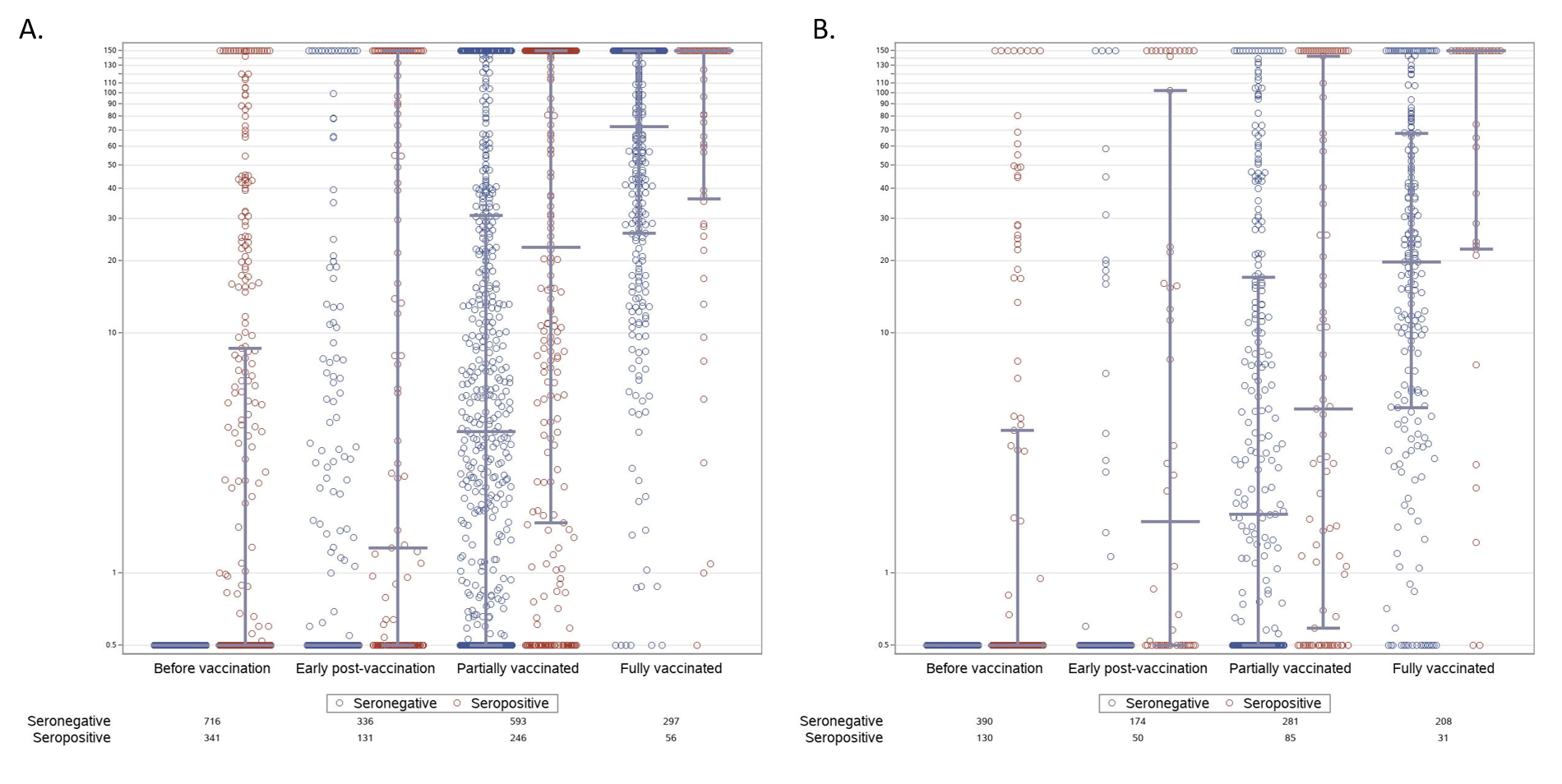
